# Coronavirus-19 and coagulopathy: A Systematic Review [COVID-COAG]

**DOI:** 10.1101/2021.01.05.20248202

**Authors:** Stephanie G Lee, Michael Fralick, Grace Tang, Brandon Tse, Lisa Baumann Kreuziger, Mary Cushman, Peter Juni, Michelle Sholzberg

## Abstract

**Background:** Understanding the association between Coronavirus Disease 2019 (COVID-19) and coagulopathy may assist clinical prognostication, and influence treatment and outcomes. We aimed to systematically describe the relationship between hemostatic laboratory parameters and important clinical outcomes among adults with COVID-19.

**Methods:** A systematic review of randomized clinical trials, observational studies and case series published in PubMed (Medline), EMBASE, and CENTRAL from December 1, 2019 to March 25, 2020. Studies of adult patients with COVID-19 that reported at least one hemostatic laboratory parameter were included.

**Results:** Data were extracted from 57 studies (N=12,050 patients) that met inclusion criteria. The average age of patients was 52 years and 45% were women. Of the included studies, 92.7% (N=38/41 studies) reported an average platelet count ≥ 150 × 10^9^/L, 68.2% (N=15/22 studies) reported an average prothrombin time (PT) between 11-14 s, 55% (N=11/20 studies) reported an average activated partial thromboplastin time (aPTT) between 25-35 s, and 34.4% (N=11/32 studies) reported a D-dimer concentration above the upper limit of normal (ULN). Eight studies (7 cohorts and 1 case series) reported hemostatic lab values for survivors versus non-survivors. Among non-survivors, D-dimer concentrations were reported in 4 studies and all reported an average above the ULN.

**Interpretation:** Most patients had a normal platelet count, elevated D-dimer, PT and aPTT values in the upper reference interval; D-dimer elevation appeared to correlate with poor outcomes. Further studies are needed to better correlate these hemostatic parameters with the risk of adverse outcomes such as thrombosis and bleeding.

## INTRODUCTION

Severe acute respiratory syndrome coronavirus 2 (SARS-CoV-2) is a new βcoronavirus that has caused the coronavirus disease 2019 (COVID-19) pandemic. Approximately 20-55% of patients hospitalized with COVID-19 have deranged hemostatic laboratory parameters suggestive of coagulopathy(1–3). These typically include increased D-dimer concentration, mildly prolonged prothrombin time (PT), mild thrombocytopenia and in late-disease, decreased fibrinogen (1,2,4–6). Available data suggest that elevated D-dimer concentration in particular is associated with poor clinical outcomes (e.g., death, transfer to the intensive care unit [ICU]). In one retrospective study, D-dimer greater than 1 µg/mL at hospital admission was associated with increased odds for death(1) while another retrospective study showed that non-survivors had higher D-dimer and longer PT, compared to survivors(4).

While coagulopathy can be associated with bleeding or clotting, available data suggests that COVID-19 is associated with a pro-thrombotic state driven by intense thrombo-inflammation likely stemming from diseased lung endothelial cells(7). In two retrospective studies, approximately one in four patients admitted to the ICU were diagnosed with venous thromboembolism (VTE)(8,9) and in one of these studies, thrombosis occurred despite thromboprophylaxis with low molecular weight heparin (LMWH)(9). In a cohort of 25 patients admitted to the ICU and screened for venous thromboembolism, 32% had confirmed pulmonary thrombosis/embolism or lower limb proximal deep vein thrombosis (DVT) despite appropriate thromboprophylaxis(10). Moreover, the median D-dimer concentration of study participants in this cohort was elevated at 2071 μg/L (range 953-3606 μg/L)(10).

The available literature has not yet been systematically reviewed and described. Such synthesis is relevant to both health care practitioners and patients in order to better understand the risk of thrombosis and bleeding that may be associated with COVID-19. The objective of our study was to systematically describe hemostatic parameters of patients with COVID-19 and their association with relevant clinical outcomes such as thrombosis, bleeding, critical illness and death.

## Methods

### Protocol and Registration

We conducted a systematic review, according to the Preferred Reporting Items for Systematic Reviews and Meta-Analyses (PRISMA) guidelines(11). Our review was registered on PROSPERO (CRD 4202017587) prior to study initiation.

### Search Strategy

PubMed (MEDLINE), EMBASE, and CENTRAL were searched from December 1, 2019 until March 25, 2020. To supplement this, Google Scholar was searched to capture grey literature and the first 100 titles were screened for eligibility. Additional articles were identified by screening reference lists of relevant articles.

A detailed search strategy guide is summarized in the Supplement. We included randomized control trials, observational studies, and case series that had at least 3 study participants. Studies of adult patients (defined as age greater than or equal to 18 years) with COVID-19 that reported at least one hemostatic parameter [i.e., platelet count, prothrombin time (PT), activated partial thromboplastin time (aPTT), fibrin degradation product (FDP), D-dimer, or fibrinogen] for the study population were included. Editorials, commentaries, letters of reply, narrative reviews, systematic reviews, meta-analyses, case reports, pediatric study populations and studies not available in the English language were excluded.

### Study Selection

Two authors (G.T. and B.T.) independently reviewed all the titles and abstracts to determine eligibility. Full-text review was performed when studies were potentially eligible for inclusion into the study. Disagreements in study inclusion were resolved through consensus, and when this failed (∼1% of studies) disagreements were resolved by a third author (S.L.).

### Data collection and quality assessment

An initial data collection tool was piloted using five articles and revised thereafter based on mutual consensus (G.T., B.T. and S.L.). The following information was independently extracted from each article by two trained investigators (G.T. and B.T.): study publication details, study design, study inclusion and exclusion criteria, study population, primary outcomes, baseline characteristics, number of study participants and hemostatic laboratory parameters (platelet count, PT, aPTT, FDP, D-dimer, and fibrinogen). Because many different d-dimer assays are clinically available, D-dimer results were reported relative to the upper limit of normal presented within each study. We also extracted details on venous thrombosis (defined as proximal venous thrombosis or pulmonary embolism confirmed by radiographic imaging), major arterial events [defined as fatal and non-fatal myocardial infarction, stroke, transient ischemic attack, arterial thromboembolism confirmed by radiographic imaging or cardiac biomarkers (troponin or creatinine kinase)], bleeding (defined as major and non-major bleeding according to the International Society of Thrombosis and Haemostasis criteria), microvascular thrombosis (defined as thrombosis of arterioles, capillaries or venules confirmed by histopathology), blood product transfusion [defined as at least one transfusion of red blood cells (RBC), whole blood, platelets, fresh frozen plasma (FFP), cryoprecipitate or fibrinogen concentrate), treatment with anticoagulation (prophylactic, intermediate or therapeutic doses) and death. When available, data were collected on hemostatic lab parameters for study participants categorized as survivors and non-survivors and as those with and without respiratory failure [defined as presence of acute respiratory distress syndrome (ARDS), need for mechanical ventilation, need for bilevel positive airway pressure (BiPAP), need for extracorporeal membrane oxygenation (ECMO) or a composite respiratory outcome as defined by study authors]. For studies that provided data for survivors and non-survivors, documentation was recorded of whether the values for survivors and non survivors were from baseline (most often date of hospitalization) or were calculated average values over the course of the hospitalization.

## RESULTS

### Study Selection & Characteristics

A total of 57 studies met inclusion criteria (see Figure 1) and a total of 12,050 patients were included in our descriptive review. Data were extracted from 1 RCT (N=199 patients), 43 cohort studies (N= 11,721 patients), and 13 case series studies (N=130 patients; Table 2).

**Figure 1.**
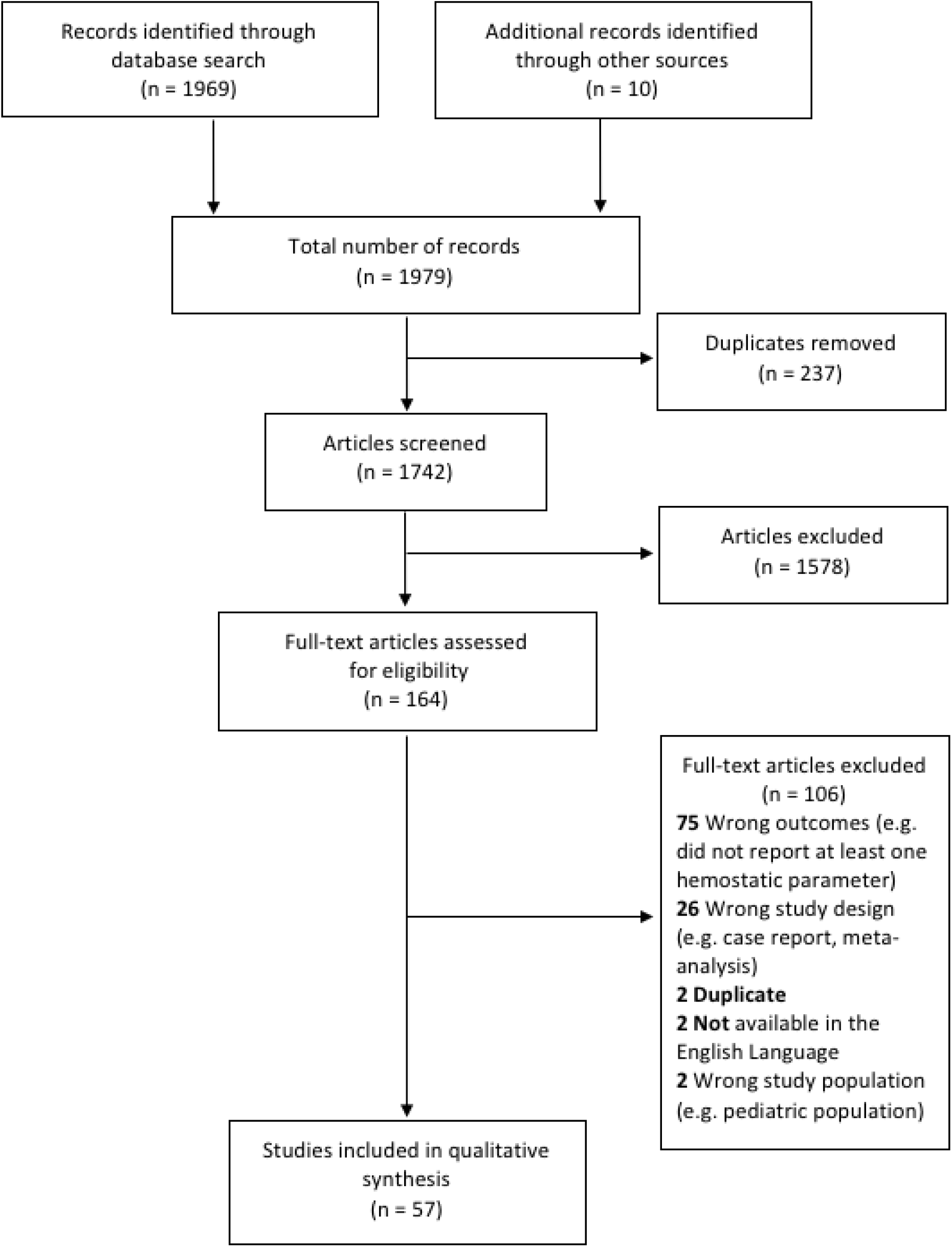
Flow diagram of included studies

Table 1 describes characteristics of the included studies. Of the 57 studies, 1 (1.8%) reported VTE outcomes, 12 (21.1%) reported major arterial event outcomes, 11 (19.3%) reported bleeding outcomes, 3 (5.3%) reported disseminated intravascular coagulation (DIC), 2 (3.5%) reported the use of anticoagulation treatment, 1 (1.8%) reported the administration of blood product transfusion and none reported on microthrombosis outcomes (Table 1).

**Table 1.**
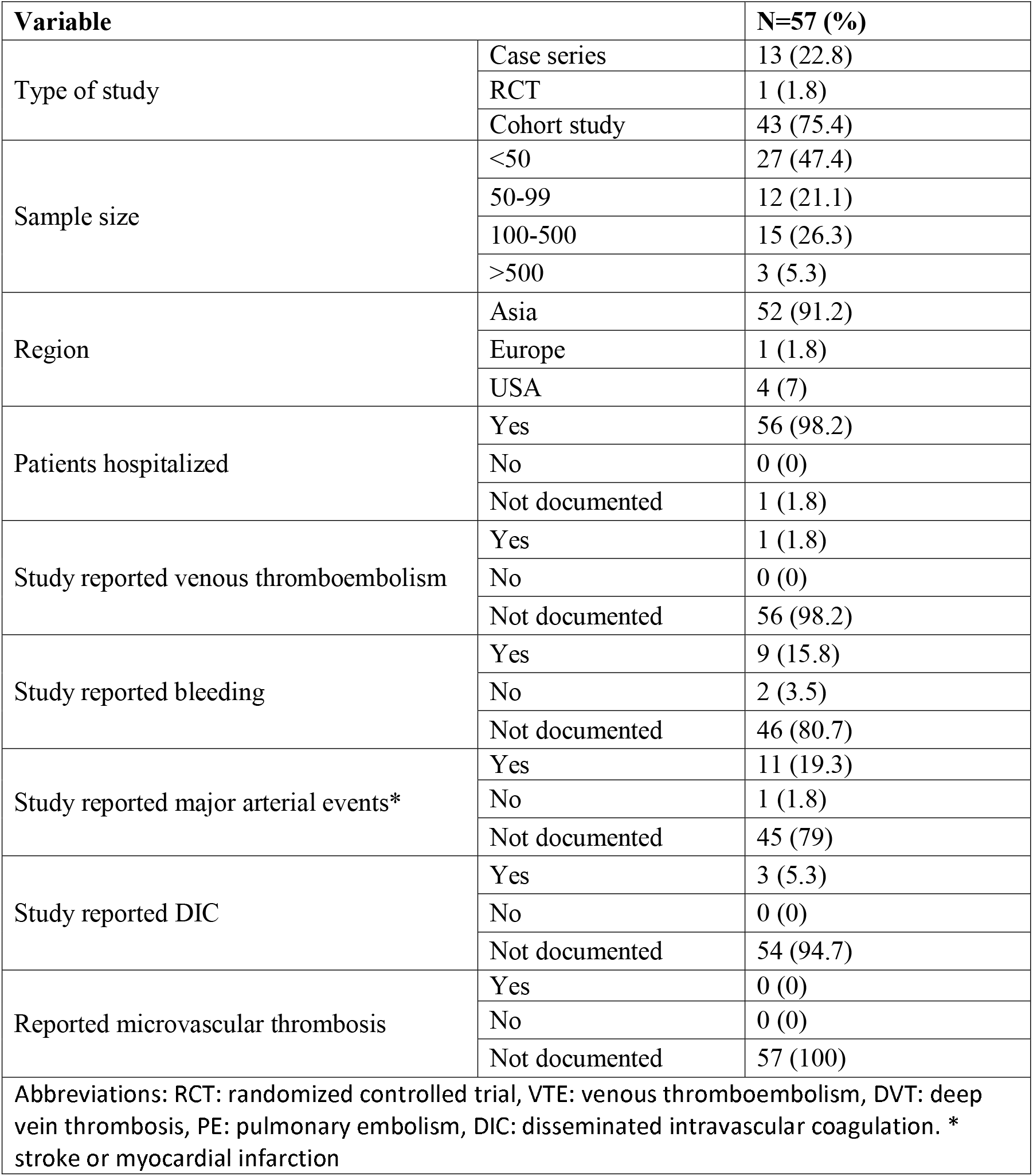
An overview of included studies.

| Variable |  | N=57 (%) |
| --- | --- | --- |
| Type of study | Case series | 13 (22.8) |
|  | RCT | 1 (1.8) |
|  | Cohort study | 43 (75.4) |
| Sample size | <50 | 27 (47.4) |
|  | 50-99 | 12 (21.1) |
|  | 100-500 | 15 (26.3) |
|  | >500 | 3 (5.3) |
| Region | Asia | 52 (91.2) |
|  | Europe | 1 (1.8) |
|  | USA | 4 (7) |
| Patients hospitalized | Yes | 56 (98.2) |
|  | No | 0 (0) |
|  | Not documented | 1 (1.8) |
| Study reported venous thromboembolism | Yes | 1 (1.8) |
|  | No | 0 (0) |
|  | Not documented | 56 (98.2) |
| Study reported bleeding | Yes | 9 (15.8) |
|  | No | 2 (3.5) |
|  | Not documented | 46 (80.7) |
| Study reported major arterial events* | Yes | 11 (19.3) |
|  | No | 1 (1.8) |
|  | Not documented | 45 (79) |
| Study reported DIC | Yes | 3 (5.3) |
|  | No | 0 (0) |
|  | Not documented | 54 (94.7) |
| Reported microvascular thrombosis | Yes | 0 (0) |
|  | No | 0 (0) |
|  | Not documented | 57 (100) |
| Abbreviations: RCT: randomized controlled trial, VTE: venous thromboembolism, DVT: deep vein thrombosis, PE: pulmonary embolism, DIC: disseminated intravascular coagulation. * stroke or myocardial infarction |  |  |

**Table 2.** Details of case series

| Author, Year | Additional Inclusion Criteria* | Country | Study Population | Sample Size | Sex, % Female | Age, years | Platelet, $\times 10^9/L$ | PT, s | aPTT, s | D-dimer | Fibrinogen, g/L | Bleeding, # of events | Major Arterial Event, # of events | Blood product transfusion, # patients received | AC given, # of patients received |
| --- | --- | --- | --- | --- | --- | --- | --- | --- | --- | --- | --- | --- | --- | --- | --- |
| Arentz, 2020 | ICU admission | USA | Hospitalized | 21 | 48 | 70 | 215 | ND | ND | ND | ND | ND | ND | ND | ND |
| Chan, 2020 | ND | China | Hospitalized | 5 | 60 | 53 | 180 | 12.8 | 38.8 | 0.5 $\mu g/mL$ | **4.68** (ULN 4.0) | ND | ND | ND | ND |
| Chang, 2020 | None | China | Hospitalized | 13 | 33 | 34 | 199 | ND | ND | ND | ND | ND | ND | ND | ND |
| Chen, 2020 | None | China | Hospitalized | 9 | 50 | 45.6 | ND | ND | ND | **0.52 $\mu g/mL$** (ULN 0.5) | **4.39** (ULN 4.0) | 0 | ND | ND | ND |
| Cheng, 2020 | None | China | Hospitalized | 11 | 27.3 | 50.4 | 143.5 | ND | ND | ND | ND | ND | ND | ND | ND |
| Ding, 2020 | Influenza | China | Hospitalized | 5 | 60 | 50.2 | ND | 13.3 | 39.8 | **0.65 mg/L** (ULN 0.5) | ND | 1 | 0 | ND | ND |
| Dong, 2020 | None | China | Hospitalized | 9 | 66.7 | 44.2 | 194.9 | ND | ND | ND | ND | ND | 1 | 1 | ND |
| Liu, 2020 | None | China | Hospitalized | 11 | 36.4 | 57.6 | 161.2 | ND | ND | ND | ND | ND | ND | ND | ND |
| Oxley, 2020 | Stroke | USA | Hospitalized | 5 | 20 | 40.4 | 297.6 | 13.8 | 31.9 | **366 ng/mL** (ULN 500) | **5.17** (ULN 4.5) | ND | 5 | ND | 2 |
| Ren, 2020 | None | China | Hospitalized | 5 | 40 | 53.6 | 189.8 | 15.3 | 42.3 | 12.69 mg/L | ND | ND | ND | ND | ND |
| Young, 2020 | None | Singapore | Hospitalized | 18 | 50 | 47 | 159 | ND | ND | ND | ND | ND | ND | ND | ND |
| Zhang, 2020 | Positive APA | China | Hospitalized | 3 | 33.3 | 68 | 112.3 | 16.4 | 45.5 | 9.02 mg/L | 5 | ND | 3 | ND | ND |
| Zhou, 2020 | Received corticosteroids | China | Hospitalized | 15 | 33.3 | 61.7 | 173.3 | 15 | 39.6 | 0.5 $\mu g/mL$ | ND | ND | ND | ND | ND |

**Note:** patients had confirmed COVID-19, except Cheng 2020, which included suspected COVID-19 cases.
Abbreviations: PT: prothrombin time, aPTT: activated partial thromboplastin time, DIC: disseminated intravascular coagulation, AC: anticoagulation, ICU: intensive care unit, ND: no data, APA: antiphospholipid antibodies

Of the 41 studies that recorded a mean or median platelet count for the study population, 38 reported an average platelet count ≥ 150 × 10^9^/L, 3 reported the average platelet count between 100-149 × 10^9^/L and none reported an average platelet count ≤ 99 × 10^9^/L. Of the 22 studies that reported a mean or median PT, 5 reported an average PT ≥14.1 s, 15 reported an average PT between 11-14 s and 2 reported an average PT ≤10.9 s. Of the 20 studies that reported mean or median aPTT, 8 reported an average aPTT ≥ 35.1 s, 11 reported average aPTT between 25-35 and 1 reported an average aPTT ≤ 24.9 s. Of the 32 studies that reported mean or median D-dimer, 11 reported an average D-dimer above the upper limit of normal (ULN), which varied between studies. Nine studies did not report the ULN; 4 of these had average D-dimer greater than 0.5 µg/mL. The mean or median of FDP was reported in 3 studies, with 2 studies reporting average FDP ≥ 10 µg/mL and 1 reporting a value of 4 µg/mL, below the reportable limit of < 5 µg/mL. The mean or median fibrinogen was calculated in 11 studies and only one of these reported an average value ≤ 0.5 g/L. The remaining 10 studies reported an average fibrinogen ≥ 2 g/L.

There were 7 cohort studies that reported hemostatic lab values for survivors (N= 1788 patients) versus non-survivors (N=420 patients). The average age of survivors was 52.3 years and of non-survivors was 66.4 years. Survivors were 37.7% women and non-survivors were 30.8% women. Of these 7 studies, the mean or median platelet count was reported in 6 studies and the average platelet count was ≥ 150 × 10^9^/L in both survivors and non-survivors. Of the non-case series that reported a mean or median value for hemostatic laboratory parameters, 92.7% (N=38/41 studies) reported an average platelet count ≥ 150 × 10^9^/L, 68.2% (N=15/22 studies) reported an average prothrombin time between 11-14 s, 55% (N=11/20 studies) reported an average aPTT between 25-35 s, and 56.3% (N=18/32 studies) There were 5 studies that reported a mean or median PT value; in those who survived, one study reported an average PT value ≥ 14.1 s, 3 studies reported an average PT value between 11-14 s and 1 study reported an average PT value ≤10.9 s. Of those who did not survive, 2 studies reported an average PT value ≥ 14.1 s and 3 studies reported an average PT value between 11-14 s. Two studies reported a mean or median aPTT value; among survivors, 1 study reported an average aPTT value ≥ 35.1 s and 1 study reported an average aPTT value between 25-35 s. In those who did not survive, 1 study reported an average aPTT value ≥ 35.1 s and 1 study reported an average aPTT value ≤ 24.9 s. There were 4 studies that reported mean or median D-dimer concentrations. In those who survived, 3/4 studies reported an average D-dimer concentration above the ULN. Among those who did not survive, all 4 studies reported an average D-dimer concentration above the ULN.

There were 3 cohort studies that reported hemostatic lab values for those with respiratory failure (N=140 patients) compared to those without respiratory failure (N=308 patients). In those with respiratory failure, 1 study reported an average platelet count ≥ 150 × 10^9^/L and 1 study reported an average platelet count between 100-149 × 10^9^/L; both studies reported an average platelet count ≥ 150 × 10^9^/L in those without respiratory failure. All three studies reported a mean or median value for PT. Among those with respiratory failure, all three studies reported an average PT value between 11-14 s and in those without respiratory failure, 1 study reported an average PT value between 11-14 s and 2 studies reported PT values ≤ 10.9 s. In those with and without respiratory value, an average aPTT value ≥ 35.1 was reported in 1 study and 2 studies reported an average aPTT value between 25-35 s. Among those with respiratory failure, 1 study reported an average D-dimer concentration below the ULN. The same study similarly reported an average D-dimer concentration below the ULN in those without respiratory failure. 1 study did not report the ULN. Among the studies that provided results from univariable or multivariable regression, elevated PT or D-dimer was consistently associated with an increased rate of worse outcomes (Table 4).

**Table 3.**
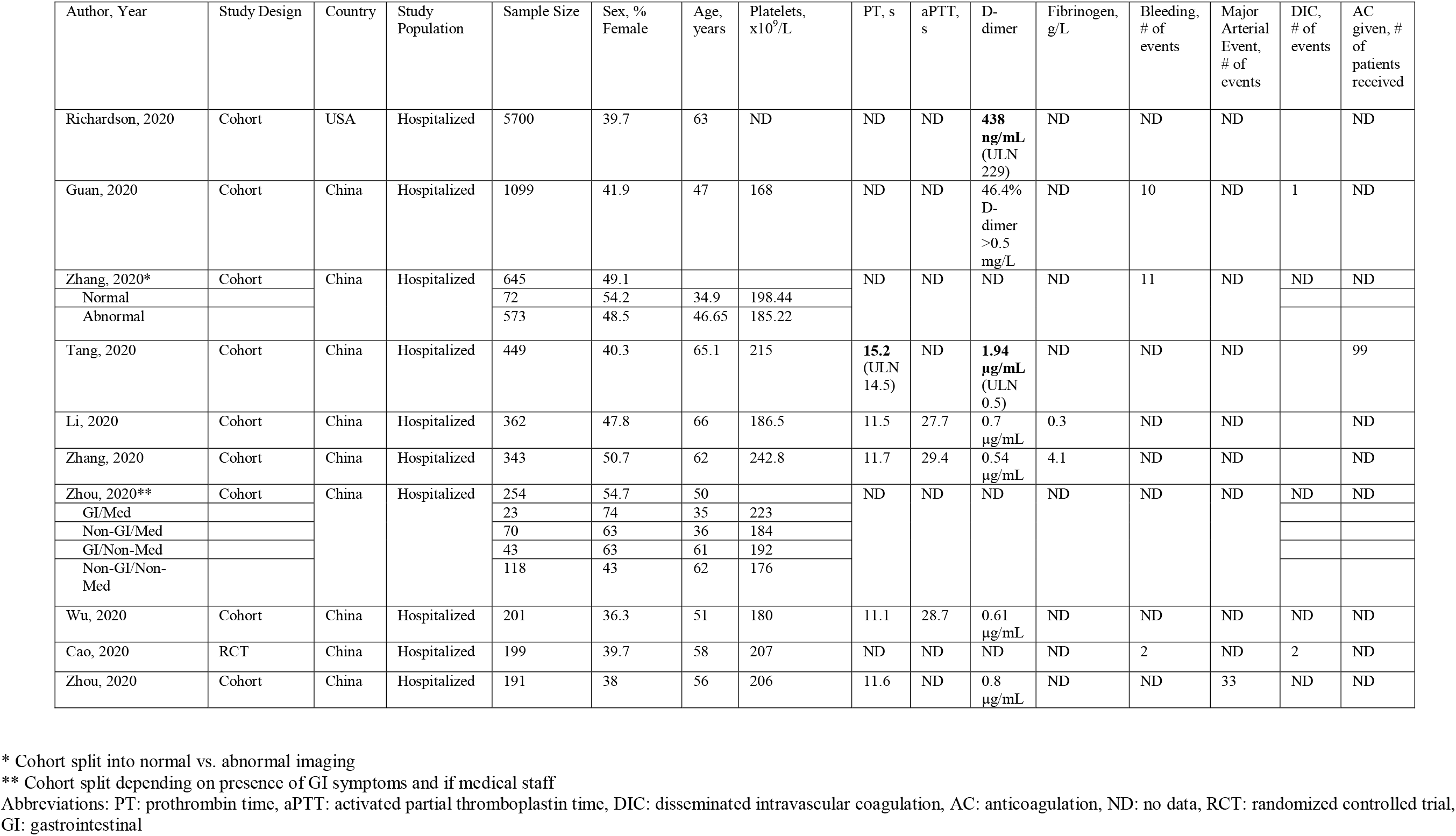
Details of 10 largest observational studies or clinical trials

| Author, Year | Study Design | Country | Study Population | Sample Size | Sex, % Female | Age, years | Platelets, $\times 10^9/L$ | PT, s | aPTT, s | D-dimer | Fibrinogen, g/L | Bleeding, # of events | Major Arterial Event, # of events | DIC, # of events | AC given, # of patients received |
| --- | --- | --- | --- | --- | --- | --- | --- | --- | --- | --- | --- | --- | --- | --- | --- |
| Richardson, 2020 | Cohort | USA | Hospitalized | 5700 | 39.7 | 63 | ND | ND | ND | **438 ng/mL** (ULN 229) | ND | ND | ND |  | ND |
| Guan, 2020 | Cohort | China | Hospitalized | 1099 | 41.9 | 47 | 168 | ND | ND | 46.4% D-dimer >0.5 mg/L | ND | 10 | ND | 1 | ND |
| Zhang, 2020* | Cohort | China | Hospitalized | 645 | 49.1 |  |  | ND | ND | ND | ND | 11 | ND | ND | ND |
| Normal |  |  |  | 72 | 54.2 | 34.9 | 198.44 |  |  |  |  |  |  |  |  |
| Abnormal |  |  |  | 573 | 48.5 | 46.65 | 185.22 |  |  |  |  |  |  |  |  |
| Tang, 2020 | Cohort | China | Hospitalized | 449 | 40.3 | 65.1 | 215 | **15.2** (ULN 14.5) | ND | **1.94 $\mu\text{g/mL}$** (ULN 0.5) | ND | ND | ND |  | 99 |
| Li, 2020 | Cohort | China | Hospitalized | 362 | 47.8 | 66 | 186.5 | 11.5 | 27.7 | 0.7 $\mu\text{g/mL}$ | 0.3 | ND | ND | | ND |
| Zhang, 2020 | Cohort | China | Hospitalized | 343 | 50.7 | 62 | 242.8 | 11.7 | 29.4 | 0.54 $\mu\text{g/mL}$ | 4.1 | ND | ND | | ND |
| Zhou, 2020** | Cohort | China | Hospitalized | 254 | 54.7 | 50 |  | ND | ND | ND | ND | ND | ND | ND | ND |
| GI/Med |  |  |  | 23 | 74 | 35 | 223 |  |  |  |  |  |  |  |  |
| Non-GI/Med |  |  |  | 70 | 63 | 36 | 184 |  |  |  |  |  |  |  |  |
| GI/Non-Med |  |  |  | 43 | 63 | 61 | 192 |  |  |  |  |  |  |  |  |
| Non-GI/Non-Med |  |  |  | 118 | 43 | 62 | 176 |  |  |  |  |  |  |  |  |
| Wu, 2020 | Cohort | China | Hospitalized | 201 | 36.3 | 51 | 180 | 11.1 | 28.7 | 0.61 $\mu\text{g/mL}$ | ND | ND | ND | ND | ND |
| Cao, 2020 | RCT | China | Hospitalized | 199 | 39.7 | 58 | 207 | ND | ND | ND | ND | 2 | ND | 2 | ND |
| Zhou, 2020 | Cohort | China | Hospitalized | 191 | 38 | 56 | 206 | 11.6 | ND | 0.8 $\mu\text{g/mL}$ | ND | ND | 33 | ND | ND |

\* Cohort split into normal vs. abnormal imaging
\*\* Cohort split depending on presence of GI symptoms and if medical staff
Abbreviations: PT: prothrombin time, aPTT: activated partial thromboplastin time, DIC: disseminated intravascular coagulation, AC: anticoagulation, ND: no data, RCT: randomized controlled trial, GI: gastrointestinal

**Table 4.** Results from studies that conducted regression analyses to determine association between coagulation parameter and clinical outcome

| Author, Year | Population | Analytic approach | Main results |
| --- | --- | --- | --- |
| Wu C, 2020 | N=201 patients with confirmed COVID-19 pneumonia admitted to Wuhan Jinyintan Hospital in China | Bivariate cox regression of factors associated with ARDS or progression from ARDS to death.<br>Laboratory parameters were included as continuous variables. | PT and D-dimer were associated with increased rate of ARDS but not aPTT.<br>D-dimer but not PT or aPTT were associated with increased rate of death. |
| Zhou F, 2020 | N=191 patients hospitalized with COVID-19 Jinyintan Hospital and Wuhan Pulmonary Hospital (Wuhan, China) | Univariable and multivariable logistic regression of factors associated with in-hospital death.<br>Laboratory parameters were transformed into categorical variables. aPTT was not included in either model. PT was included only in the univariable model. | Univariable model<br>PT ⇒ 16 was associated with increased odds of death (OR 4.62, 95% CI 1.29,16.50).<br>D-dimer $> 1$ ug/mL was associated with increased odds of death (OR=20.04, 95% CI 6.52,61.56)<br>Multivariable model<br>D-dimer $> 1$ ug/mL was associated with increased odds of death (OR=18.42, 95% CI 2.64,128.55) |
| Mo, 2020 | N=155 consecutive patients with confirmed COVID-19 in Zhongnan Hospital of Wuhan University | Multivariable regression of factors associated with refractory COVID-19 pneumonia<br>Platelet count was the only coagulation parameter included | Platelet count was not associated with an increased odds of refractory pneumonia. |
| Liu, 2020 | N = 78 patients hospitalized at three tertiary care hospitals in Wuhan | Univariable and multivariable regression of factors associated with progression of COVID-19.<br>Only d-dimer and platelet count were included (both as categorical variables) and results were only provided for univariable model. | Univariable model<br>For odds of progression of COVID-19: Platelet count $< 100 \times 10^9$ L OR = 2.26 (95% CI 0.39-12.96), D-dimer $> 1$ ug/mL OR = 1.80 (95% CI 0.36-8.93), though both had wide confidence intervals including possibility of null effect |
| Gao, 2020 | N=43 patients hospitalized with COVID-19 in Fuyang Second people's hospital | Complete details were lacking, but the authors created a logistic regression model to identify factors associated with severe COVID-19<br>Only D-dimer was included and it was included as a categorical variable | D-dimer $> 0.28$ μg/L was associated with an increased odds of severe COVID-19 (OR=12.14(95% CI 1.72,85.86) |
| Tang, 2020 | N=449 patients with severe COVID-19 admitted to Tongji Hospital of Huazhong University | Multivariable regression of factors associated with 28-day mortality.<br>D-dimer, PT, and platelet values were included as continuous variables | Higher D-dimer and PT values were associated with an increased odds of mortality. Higher platelet count was associated with lower odds of mortality. |
| Zhang, 2020 | N=343 patients hospitalized at the Wuhan Asia General Hospital | Multivariable Cox proportional hazards model. Details were lacking on the included variables, though the manuscript suggests that D-dimer was the only coagulation parameter included. | D-dimer levels $\geq 2.0$ μg/ml was associated with higher rate of death compared to adults with a value below 2.0 ug/mL (HR=22.4, 95% CI 2.86-175.7 |

Legend: OR =odds ratio, HR = hazard ratio, PT = prothrombin time, aPTT = activated prothrombin time

Of the 57 studies that reported hemostatic laboratory parameters, one study (N=20 patients) reported venous thrombosis and 12 studies reported major arterial events. Of these 12 studies, 11 studies (N=106 patients) documented a major arterial event and 1 study documented the absence of major arterial events. The etiology of major arterial events was defined as elevated cardiac biomarkers in 6/11 studies (N=83 patients), myocardial infarction/arrest in 2/11 studies (N=5 patients), ischemic stroke in 2/11 studies (N=8 patients) and 1 study documented acute cardiac injury (N=10 patients). There were 12 studies that reported bleeding outcomes where 9/12 studies (N=37 patients) recorded bleeding as an adverse outcome and 3/12 studies documented the absence of bleeding as the absence of an adverse outcome. The cause of bleeding was hemoptysis in 7/9 studies (N=33 patients) and gastrointestinal (GI) bleeding in 2/9 studies (N= 4 patients). The remaining studies did not comment on the presence or absence of thrombotic or bleeding events.

## DISCUSSION

In this systematic review of 57 studies including over 12 000 patients, most patients hospitalized with COVID-19 had a normal platelet count, near normal PT and PTT values and elevated D-dimer concentrations. There were limited data on how these hemostatic parameters were associated with clinical outcomes, although elevated D-dimer concentration appeared to be the strongest predictor of worse clinical outcomes.

These results provide a framework to guide clinicians caring for patients with COVID-19. For example, we observed that thrombocytopenia is uncommon among patients hospitalized with COVID-19 and such a finding should prompt clinicians to search for alternate causes of thrombocytopenia. The reason for near normal platelet counts in hospitalized patients with COVID-19 is unknown but could potentially be due to inflammatory thrombocytosis related to increased interleukin-6 (IL-6) levels that occur in COVID-19(1,7,12–14). Our study also supports the observation that elevated D-dimer is associated with poor clinical outcomes. Data from hospitalized patients in Wuhan identified that patients requiring critical care had a median D-dimer concentration of 2.4 mg/L compared to 0.5 mg/L in those not requiring critical care(6) and a second study in New York showed that a D-dimer concentration of 4-times above normal was associated with an approximately 5-fold higher odds of critical illness compared to normal a D-dimer concentration(15). This information can help physicians triage patients with COVID-19 to the appropriate disposition and supports the recommendation that inpatients with COVID-19 that have elevated D-dimer concentrations need to be monitored closely(16). Another important question is whether patients should be anticoagulated based on elevated D-dimer. In a retrospective study of 449 hospitalized patients with severe COVID-19, patients with D-dimer >6-times above normal who received heparin thromboprophylaxis (mostly enoxaparin 40-60 mg/day) had an approximate 20% lower mortality compared to those who did not receive thromboprophylaxis(17). Clinical trials are currently ongoing to understand whether these patients might benefit from higher doses of anticoagulation.

It has been hypothesized that COVID-19 associated coagulopathy is driven by pulmonary vascular endotheliopathy, which may contribute to the risk of thromboembolism and death in hospitalized patients with COVID-19. Whether this endotheliopathy is due to direct effects of the virus or secondary to immune activation and diffuse inflammation remains unknown (7,12,18). Although DIC also involves endothelial cell activation, it appears mechanistically different from COVID-19 associated coagulopathy in that the latter is predominantly prothrombotic and the underlying inflammatory stimulus typically does not lead to consumption of hemostatic elements(7).

There are a number of important limitations to our study. First, most of the included patients were from China and thus the rates of thromboembolism might be underestimated as Asian ethnicity appears to be associated with a lower risk of thromboembolism(19). Second, most included studies did not quantify the association between abnormal coagulation parameters and the risk of adverse events (e.g., respiratory failure, mortality) and those that did generally lacked external validation. Third, there was high variability in the type of D-dimer assay used across studies, which made it inappropriate to meta-analyze results. Fourth, we only studied hospitalized adult patients, so it is unknown how results apply to children or other patient populations such as ambulatory patients or pregnant women.

Necessary future directions include antithrombotic clinical trials in the pre-hospital, pre-intensive care, intensive care, and post-hospital care settings. Clinical trial emphasis on speed and pragmatism is required given the immediate clinical need and risk of loss of equipoise. Furthermore, prediction models are needed to understand which hemostatic laboratory parameters, or combination thereof, portend the highest risk for adverse outcomes.

Our summarized data provide a systematic review of the literature describing COVID-19 associated coagulopathy, the thrombo-inflammatory pattern of COVID-19 associated coagulopathy, and its association with poor clinical outcomes. This information will help frontline health practitioners use hemostatic laboratory parameters for prognostication as well as the triage of patients to the appropriate disposition, which is particularly crucial during a pandemic if resources become depleted.

## Data Availability

N/A

